# Long-Term Impact of Stressful Life Events on Breast Cancer Risk: A 36-Year Genetically Informed Prospective Study in the Finnish Twin Cohort

**DOI:** 10.1101/2024.08.27.24312571

**Authors:** Elissar Azzi, Hannes Bode, Teemu Palviainen, Mikaela Hukkanen, Miina Ollikainen, Jaakko Kaprio

## Abstract

Breast cancer (BC) is influenced by both genetic and environmental factors, but the long-term impact of stressful life events (SLEs) remains unclear. We examine the association between SLEs and BC risk using cohort and twin-pair analyses with 36 years of follow-up in the Finnish Twin Cohort, including 10,342 women and 719 BC cases. SLEs were assessed in 1981 by a questionnaire, while cancer incidence and mortality data were obtained from Finnish registries. Polygenic risk score for breast cancer (PRS-BC) and DNA methylation (DNAm) profiling were used to explore the underlying genetic and epigenetic factors. Cox proportional hazards models showed a significant association between SLEs and breast cancer risk (HR = 1.05 per event, 95% CI 1.02-1.08). As few as 2-3 SLEs were associated with a 24% increased risk (HR = 1.24, 95% CI 1.00-1.54), emphasizing the impact of even a modest number of events. Within-pair analyses in monozygotic twins suggested non-genetic factors mediate this association. Stratification by birth cohort revealed a stronger effect in women born before 1950 (HR = 1.07, 95% CI 1.01-1.12). While PRS-BC was not significantly associated with breast cancer risk, DNAm analysis identified 42 BC-associated CpG sites linked to both SLE exposure and environmental BC risk. These findings were replicated in cancer-free twin pairs, supporting epigenetic rather than genetic mediation. SLEs may be an independent risk factor for breast cancer, potentially mediated by epigenetic mechanisms. Further research is needed to explore the functional consequences of stress-related epigenetic changes and their role in BC development across generations.

## INTRODUCTION

Breast cancer remains the predominant and lethal form of malignancy affecting women worldwide, with 2.3 million new cases diagnosed globally in 2020 alone ^1^. Twin studies reveal that 31% of variability in risk is attributed to genetic factors ^2^, encompassing well-known high-impact genes like *BRCA1/2*, *CHECK2*, and *PALB2* ^3^, as well as numerous genes with single nucleotide polymorphisms (SNPs), collectively quantifiable in a polygenic risk score (PRS) ^4^. While significant progress has been made in understanding the genetic underpinnings of breast cancer, the full picture of its aetiology remains a work in progress. Environmental factors and health-related behaviours, including established ones such as alcohol consumption, obesity, female reproductive factors, and hormone exposures, contribute significantly to breast cancer risk ^5^. However, the role of psychological stress, particularly in the form of stressful life events (SLE), remains an area of ongoing debate in breast cancer epidemiology.

Stress – the physical and emotional response to challenging or demanding situations – has potential implications for later life health through various physiological mechanisms. Chronic or repeated stress is hypothesized to contribute to tumorigenesis through physiological mechanisms such as immune suppression, endocrine dysregulation, and inflammatory processes ^6^. These disruptions are thought to occur through prolonged activation of the hypothalamic-pituitary-adrenal (HPA) axis, which can lead to elevated cortisol levels ^6^. Cortisol is a hormone essential for mammary gland function and has been implicated in promoting cell proliferation and tumour growth ^7^. Chronically elevated cortisol impedes the immune system, hindering its ability to eliminate abnormal cells and fostering a pro-inflammatory environment ^8^. Emerging evidence suggests that stress-related biological effects may also be mediated by epigenetic mechanisms, particularly DNA methylation ^9–11^. This can lead to persistent changes in gene expression relevant to cancer development and may be reflected in epigenetic profiles ^12^.

The link between stressful life events and breast cancer risk is a complex one, with ongoing epidemiological research exploring the potential connection yielding diverse results. While some studies suggest a positive association between stressful life events and breast cancer risk ^13^, others, such as the UK study of 106,000 women ^14^ report no consistent evidence for an association of breast cancer risk with perceived stress levels or adverse life events in the preceding 5 years. A 2019 meta-analysis assessing the impact of stress, or lack thereof, on breast cancer incidence, provided a broader perspective highlighting a moderate overall association between stressful life events and breast cancer risk [(pooled risk ratio: 1.11 (95%CI 1.03-1.19)] ^15^. A prior study using the same Finnish Twin Cohort demonstrated an association between stressful life events and increased breast cancer incidence ^16^. However, the study’s 15-year follow-up period provided a limited window to explore the long-term effects of stress on breast cancer risk. Additionally, the underlying mechanisms behind this association—whether it is driven by shared genetic predisposition, environmental stressors, or epigenetic factors—remain unclear.

To address these gaps, our study extends the follow-up period to 36 years and incorporates genetic and epigenetic analyses to better understand the relationship between stress and breast cancer. Specifically, we assess polygenic risk scores (PRS) to evaluate genetic predisposition and analyse DNA methylation (DNAm) patterns to explore potential epigenetic mechanisms. PRS for breast cancer (PRS-BC), derived from numerous breast cancer-associated low-impact genetic variants identified in genome-wide association studies, helps estimate inherent susceptibility to breast cancer and provides insights into whether genetic factors contribute to the observed association with stressful life events.

Meanwhile, DNA methylation serves as a dynamic epigenetic modification that may mediate the link between exposure and breast cancer risk by altering gene expression in ways that promote tumorigenesis. Environmental exposures, including stress, have been shown to induce significant epigenetic changes, such as altered patterns that may contribute to breast cancer risk. In support of this, Bode et al. (2024) identified 212 with overall environmental risk for breast cancer, detectable on average 11 years before diagnosis in Finnish twin pairs discordant for breast cancer diagnosis ^16^. These findings highlight the potential role of epigenetic mechanisms in linking external stressors to tumour development, providing a foundation for our investigation into how stressful life events influence breast cancer risk through long-term epigenetic modifications.

Our investigation employs both standard cohort analyses and within-pair comparisons of monozygotic (MZ) and dizygotic (DZ) twin pairs, concordant and discordant for breast cancer. Twin pairs provide a natural control for shared genetic and early environmental factors, allowing us to separate genetic influences from environmental effects. By comparing breast cancer risk within twin pairs discordant for stressful life event exposure, we can assess the impact of stress while accounting for familial influences. Within-pair models were analysed separately for MZ and DZ pairs to determine whether genetic similarity modified the association between stress and breast cancer incidence. Additionally, we stratify analyses by birth decade to account for temporal variations in environmental exposures. By integrating genetic data, epigenetic markers, and detailed records of stressful life events, this study provides a more comprehensive understanding of how stress may contribute to breast cancer risk over the lifespan.

## METHODS

### Stressful Life Events Analysis

#### Twin cohort description

The data used in this study originates from the older Finnish twin cohort, consisting of same-sex twin pairs born before 1958 ^17^. This cohort was followed up with comprehensive health and lifestyle questionnaires in 1975, 1981, 1990 and 2011. Of the 16160 female twins in the cohort, 12986 returned a questionnaire in the 1981 survey. The questionnaire was mailed in the autumn of 1981, with reminders into early 1982. The first questionnaires were returned on October 7, 1981, with a median return date of November 4, 1981. Follow-up for each person started from the date of return of the questionnaire and ended on December 31, 2018. After excluding those individuals with prevalent BC or residence outside Finland, missing life event information and other covariates, final sample size was 10342, with 719 incident BC cases from 1982 to 2018.

#### Risk factors

SLE data from the past 5 years were primarily collected via the 1981 questionnaire. Follow-up for BC incidence extended from 1982 to 2018, with cancer diagnoses tracked using the Finnish Cancer Registry. The compilation of data on SLEs is explained in more detail by Lillberg et al. (2003) ^16^. Essentially, the 1981 questionnaire contained questions on 17 major life events, including change of residence, loss of job, death of a relative and others (Supplementary list 1). These were aggregated for analysis on an individual basis as a SLE score, with the number of events exceeding 10 reduced to 10, as persons with more than 10 events were few in number. Other risk factors of interest were compiled from the 1981 questionnaire but supplemented with 1975 information where missing. The covariates were zygosity, marital status, oral contraceptive use, social class, BMI (calculated as the mean between 1975 and 1981), alcohol consumption (expressed in ethanol grams per day and categorised as abstainer, 1-10, 11-20 and 21 or more grams per day), smoking (never, former, current), and leisure time physical activity (quintiles, MET-hours per week). In addition, we assessed the link between BC and reproductive history using latent classes of lifelong pregnancy data obtained from the 1975 questionnaire and the Finnish Population Register (hereafter “reproductive class” described in Hukkanen et al., 2024) ^18^.

#### Outcome

Data on cancer incidence up to the end of 2018 was obtained from the Finnish Cancer Registry (FinData permit THL/6353/14.06.00/2023), while data on mortality and emigration were obtained from the Finnish Population Register and Statistics Finland ^17^. Cases were defined as women who did not have BC prior to response to the 1981 questionnaire but were diagnosed with BC thereafter by the end of 2018. In contrast, controls were defined as individuals who were not diagnosed with BC until the time of censoring, due to emigration, death or end of the study in 2018.

#### Evaluating the association of stressful life events with breast cancer risk using polygenic risk scores

Genotypes for evaluating genetic BC cancer risk through PRS-BC was available for a sub-sample of 4,601 women (4,237 controls and 364 cases, disease prevalence 7.9%). Within this sample, 176 DZ twin pairs were discordant for BC. The PRS-BC was constructed using 1,069,529 single nucleotide polymorphisms (SNPs) ^4^ identified from a large genome-wide association study (GWAS) ^19^ on BC. PRS-BC was calculated as a weighted sum of BC risk alleles using effect sizes from the GWAS summary statistics. For details see Supplemental Material: Genetic Analyses.

#### Evaluating the association of stressful life events with breast cancer risk using DNA methylation

Altogether 319 twin pairs (172 MZ and 147 DZ pairs) had information on DNAm data for 212 CpG sites related to BC risk and the 1981 SLE score. DNAm, was generated by Illumina Infinium MethylationEPIC (EPIC) platform (Illumina, San Diego, CA, USA). Quality control and preprocessing of the data followed our in-house pipeline ^20^. Bode et al. (2024) ^20^ identified 212 CpG sites in genome-wide blood DNAm data associated with future BC risk, based on a discordant twin pair design in which one twin sister in each pair developed BC later and the other twin sister was unaffected. This approach controls for technical confounding factors, age and shared environment, as well as germline genetics ^21^. These results provide support for the view that the identified CpG sites’ associated with BC risk in survival analysis likely reflects within pair differences in environmental exposures that impact BC risk ^20^. For details see Supplemental Material: DNA Methylation Analyses. To avoid confounding by BC or related factors, twin pairs where either of the twins had a BC diagnosis before DNA sampling were excluded. No exclusions were made based on other diseases occurring before or after DNA sampling.

#### Statistical Analysis

Cox proportional hazards survival model was used to estimate the risk of BC incidence according to the number of SLEs. The number of SLEs was modelled both as a continuous variable and as an ordinal variable in six categories (0-1, 2-3, 4-5, 6-7, 8-9 and 10 or more) and estimated the risk for each category to allow for non-linear effects. Cox proportional hazards models were analysed adjusting for age (as the time-scale variable), with follow-up from birth, but entry into the model at the time of response to the 1981 questionnaire, until death, emigration or end-of-follow-up on December 31, 2018. Initial models were adjusted for age while multivariable models were additionally adjusted for potential BC covariates listed above. Results from the models are reported with all covariates included. The effect of each of the covariates (other than age or sex) on the risk of BC was also examined (data not shown). Within-pair Cox-regression models of life events and mortality were performed with the baseline hazard specific to each pair. The models yield overall estimates adjusted for familial and genetic factors shared by the twins in a pair. The proportional hazards assumption of the Cox regression model was examined by graphical methods of plotting log–log survival curves and tested using a global test based on Schoenfeld residuals.

Statistical analyses were done using Stata (v18, Statacorp, College Station, TX, USA). After obtaining our primary results, we ran sensitivity analyses examining the effect of birth cohort, in which our focus was on the effect sizes and not statistical significance testing. We report the means and standard deviations of continuously distributed covariates and the distributions of categorical covariates.

Linear models were used to test for association between PRS-BC and SLE scores. Cox regression models were used to examine the association between PRS-BC and BC among all women with genetic data, and within DZ pairs discordant for BC. Linear models were employed to analyse the association between within pair difference in DNAm at 212 selected CpG sites ^20^ and the within pair difference in SLE exposure. Two models were used, one using all available twin pairs and the other examining MZ pairs alone to fully control for genetic effects. Further, we examined whether the effect of SLEs on DNAm aligned with the direction of DNAm associated with environmental BC risk as reported in Bode et al. (2024) ^20^. Each CpG site with the same effect direction as in Bode et al. (2024) ^20^ and p<0.05 were considered as validated.

## RESULTS

### Stressful Life Events and Breast Cancer Incidence

#### Cohort analyses

During the 36-year follow-up, 719 incident BC cases were recorded in our cohort. Participating women were aged 24 years and older in 1981 with a mean age of 39 (SD 12) years; 32% were MZ twins. The cumulative hazard by age is shown in Supplemental Figure 1, with an overall cumulative incidence of 7%. The distribution of baseline characteristics in cases and controls is detailed in Supplemental Table 1.

The relationship between the number of SLE and BC risk is shown in Table 1, whereby accumulating numbers of SLE increases the risk of BC incidence. After adjusting for age, the hazard ratio for BC per one-event increase in the total number of life events was 1.05 (95%CI 1.02-1.08) and remained consistent at 1.04 (95%CI 1.00-1.07) after adjusting for additional potential confounders (zygosity, marital status, oral contraceptive use, social class, reproductive class, BMI, alcohol consumption, smoking, and leisure time physical activity). Being exposed to 10 or more SLE increased the risk of BC incidence by 93% (Table 1). Within-pair analysis showed that for every additional SLE, MZ pairs had a higher within-pair HR than DZ pairs (1.14 [95%CI 1.01-1.29] vs 1.03 [95%CI 0.95-1.11]) for BC, indicating a significant risk increase even after controlling for genetic background. The individual based analyses resulted in HRs with the same magnitude as the within DZ pair analysis (Table 1). The impact of known and proposed BC risk factors on HRs for BC incidence are detailed in Supplemental Table 2.

**Table 1.**
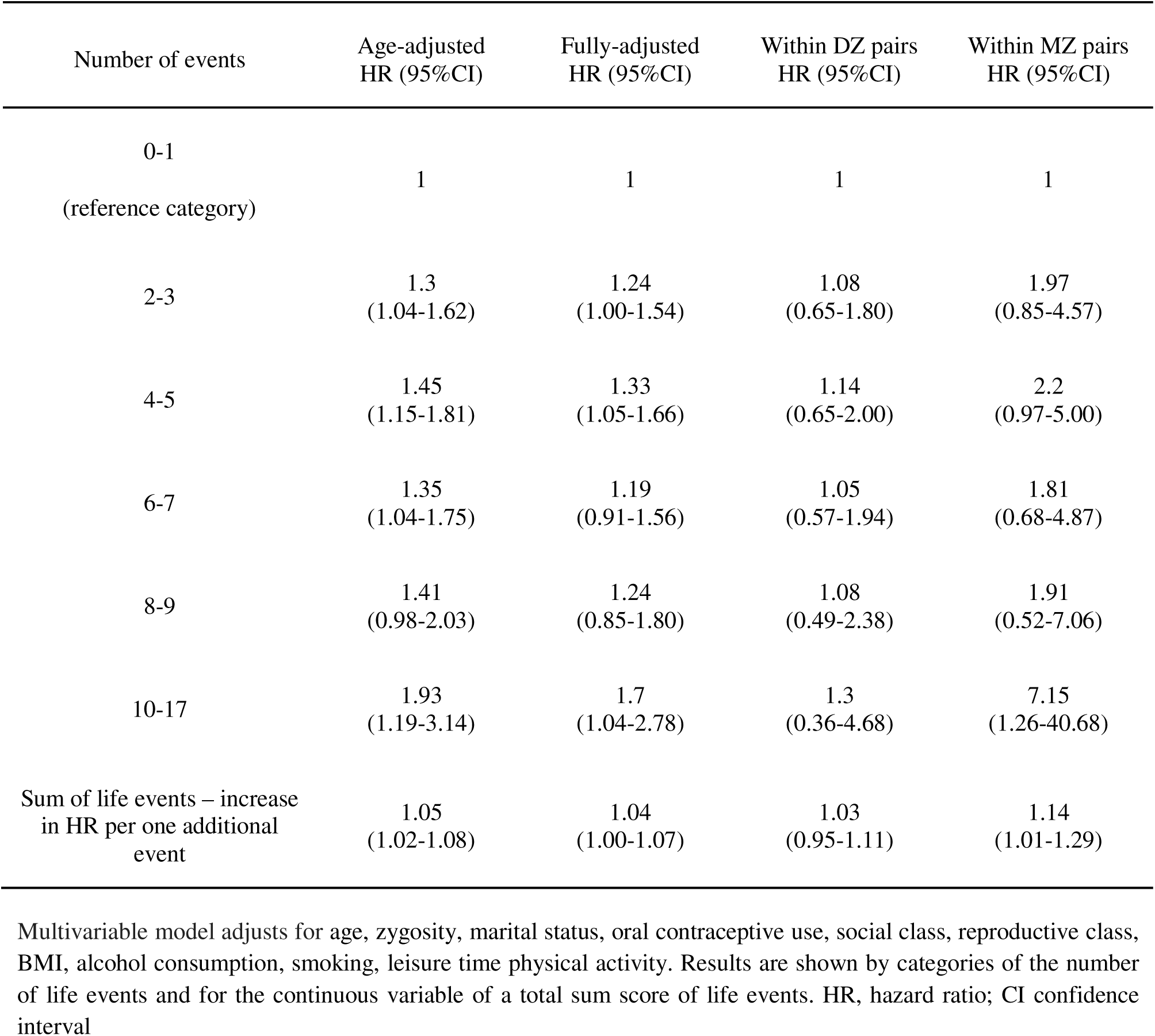
Survival model estimates of hazard ratios associated with the number of stressful life events and every one event increase in the sum of reported life events in 1981 and breast cancer incidence between 1982 and 2018 in all individuals and within MZ and DZ pairs.

#### Stratification by birth decade

The extensive range of birth years in the cohort enabled the implementation of a segmented analysis to examine the impact of SLEs across birth decades. Stratifying by birth decade revealed a modest increase in HR for those born before 1950 compared to those born after 1950 (HR 1.07 vs.1.01, Table 2, Supplemental Table 3). A deeper look into the other BC risk factors uncovered some additional temporal differences: The most notable difference was the HR associated with use of oral contraceptives in those born before 1950 (HR 1.28; 95%CI 1.03-1.59) being appreciably higher than that of those born after 1950 (HR 1.01; 95%CI 0.75-1.35). Of other factors, being single and alcohol consumption exhibited modest variations when segregated by birth cohort (Supplemental Table 3).

**Table 2.** Hazard ratios for SLE score by birth cohort segments.

| Birth cohort | All | Before<br>1940 | 1940-1949 | 1950-1957 |
| --- | --- | --- | --- | --- |
| Age at response to 1981 survey<br>(min, max) |  | 41.8, 92.2 | 31.8, 43.2 | 23.8, 33.4 |
| SLE score –<br>increase in HR per one additional event<br>(95%CI) | 1.05<br>(1.02-1.08) | 1.04<br>(0.98-1.11) | 1.07<br>(1.01-1.12) | 1.01<br>(0.96-1.06) |
| Number of breast cancer cases | 719 | 222 | 230 | 267 |
| Number of women | 10342 | 3349 | 3049 | 3944 |
| HR, Hazard ratio; CI, Confidence interval |  |  |  |  |

### Genetic breast cancer risk, cancer incidence and association with stressful life events

Among women with a PRS-BC available, there were 364 BC cases, while 4237 women were unaffected by the end of the 36-year follow-up. Adjusting for underlying family structure, we observed a 3% increase in BC risk per SD increase in BC PRS, with a HR of 1.03 (95%CI 0.92– 1.15). Among 176 DZ twin sister pairs discordant for BC diagnosis, a pairwise survival model yielded an HR of 1.03 (95%CI 0.89–1.20). For all women with available data (n=3906), the correlation between SLEs and BC PRS was -0.01 (95%CI -0.04 – 0.02).

### Environmental Breast Cancer Risk related DNA methylation

To assess if environmental BC risk related DNAm is associated with SLEs, we followed up our previous findings on 212 such CpG sites (Bode et al., 2024) ^20^ in the 319 pairs with SLE score and sampled for DNAm analysis, on average, 16 years prior to BC diagnosis. Their mean age at the 1981 questionnaire study was 40.0 (SD 9.8) years, and mean age at DNA sampling was 63.2 (8.9) years; these ages did not differ by zygosity. The intraclass correlation for the SLE score was 0.44 (95%CI 0.31–0.55) in MZ pairs indicating substantial non-genetic influence.

Our analysis implicated 42 of these previously identified CpG sites, with significant associations between SLE exposure and BC risk related DNAm. These CpG sites are in 32 known genes (Supplemental File 1). In total, 197/212 CpG sites displayed association trend with SLEs in the same direction as with BC risk reported by Bode et al. (2024) ^20^ (Supplemental File 1). This significant skew towards hypomethylation associated with SLE exposure (binomial test p = 1.19*10^-41^ is in line with our previous findings that showed that hypomethylation associates with an exposure to environmental BC risk factors (Bode et al., 2024) ^20^ (Figure 1).

**Fig. 1.**
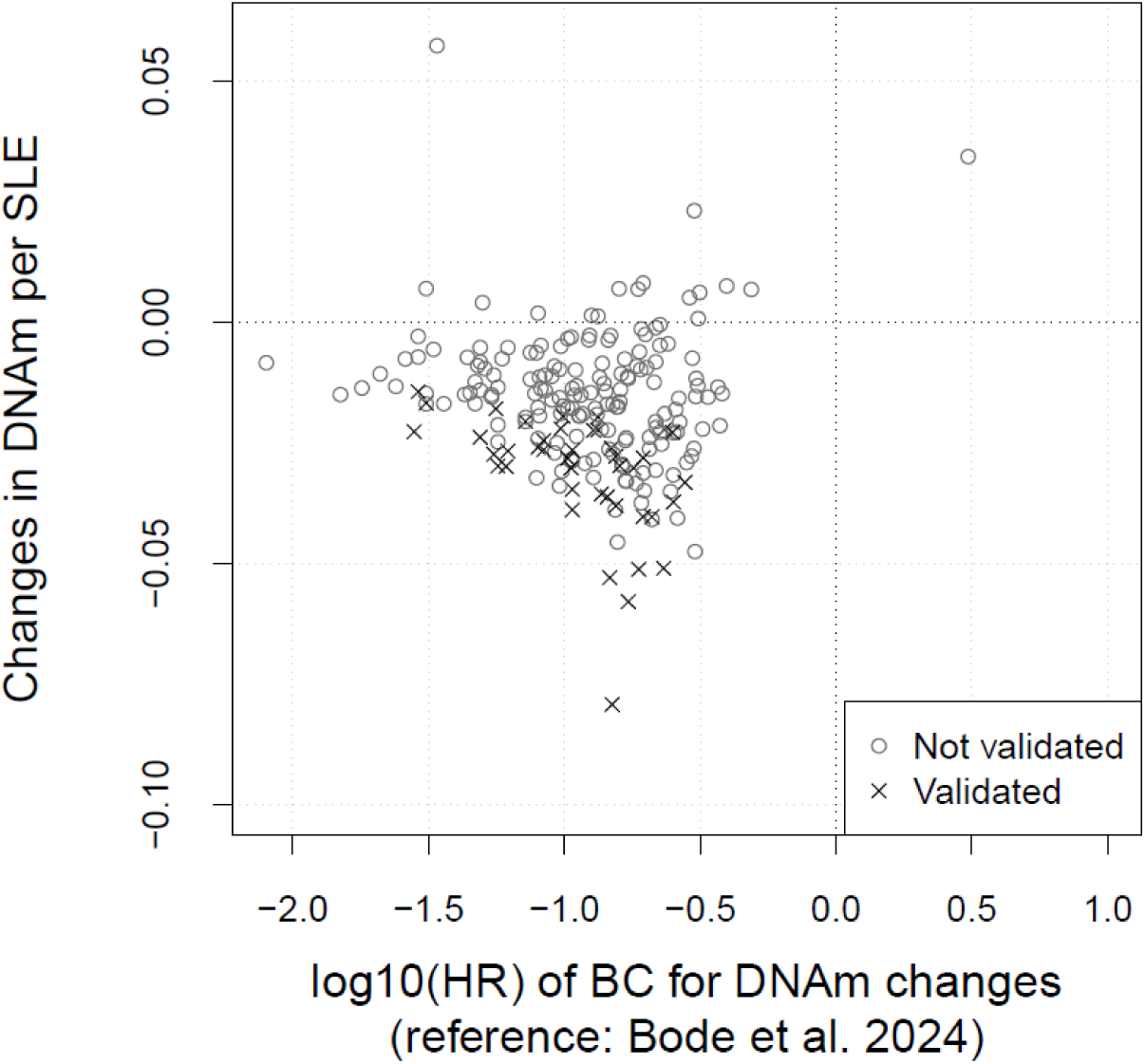
Association between stressful life events exposure and DNAm differences among all pairs. On the y-axis the effects of exposure to stressful life events (SLEs) on BC related differences in DNAm in twin pairs (172 DZ and 147 MZ pairs) for 212 CpG sites identified in Bode et al. (2024) ^20^. On the x-axis the hazard ratios (HR) of breast cancer (BC) in relation to changes in DNAm according to Bode et al. (2024) ^20^. The validated data points indicate that the association between SLEs and DNAm have a p-value < 0.05.

To further isolate the influence of germline genetics on the observed effects of SLEs on DNAm, we repeated the analysis solely on the 174 MZ pairs across all 212 DNAm sites (Supplemental File 1). The effect sizes of the within pair difference in SLEs on the within pair difference in DNAm at the aforementioned 42 CpG sites were 36% larger in MZ pairs compared to DZ pairs (paired t-test, p = 0.012). This suggests that the observed associations between exposure to SLEs and DNAm are largely independent of germline genetic factors. Further, within the MZ twin pairs the effect direction in 193/212 CpG sites was in line with that observed in Bode et al. (2024) ^20^ for BC risk (binomial-test p = 1.9*10^-37^).

As a sensitivity test, we repeated the analyses in the Older Finnish Twin Cohort with 154 female twin pairs (96 MZ and 58 DZ pairs) who remained cancer-free until the end of the follow-up in 2018, to exclude the potential effect of future cancer diagnosis on the association between DNAm at the 212 CpG sites and SLEs. A total of 174 of the 212 CpG sites (binomial test, p = 1.1*10^-20^) showed directional consistency with Bode et al. (2024) ^20^ (Supplemental Table 1). We further restricted the analysis to the 96 MZ twin pairs (Supplemental Table 1), leveraging their matched germline genetics despite the smaller sample size. Even within this subset, 182 out of 212 CpG sites showed directional consistency with Bode et al. (2024) ^20^ (binomial test, p = 9.8*10^-28^). These findings strengthen the evidence that DNAm patterns associated with environmental BC risk might also be linked to SLEs exposure, independent of both germline genetics and future BC diagnosis.

## DISCUSSION

We conducted a genetically informed study to investigate the long-term impact of stressful life events on breast cancer incidence over a 36-years of follow-up period. Our results underscore a significant association between cumulative exposure to stressful life events and breast cancer risk while pointing towards the limited role of genetic risk. Instead, our study suggests that DNA methylation could play a role in mediating the effects of stressful life events on breast cancer incidence. Intriguingly, our analysis also revealed a birth-decade difference in risk factor association, with a higher hazard ratio for individuals born before 1950, indicating that the impact of stressful life events on breast cancer risk may vary across generations.

It is well established that the balance of neuroendocrine hormones in women is easily affected by psychological trauma ^22^, though the impact of stressful life events on health outcomes, especially cancer, is far from fully understood. Major traumatic events are potent sources of stress and more easily attributable to adverse health events later in life ^23^. In contrast, less evidently traumatic events and daily stressor in people’s lives may be far more insidious in their long-term biological consequences. A previous study utilizing the Finnish Twin Cohort ^23^showed a heightened association between accumulation of stressful life events, in particular of those of greatest a-priori-interest, such as divorce or death of a loved one, and increased risk of breast cancer incidence during 15 years of follow-up ^16^. Our current study extends the follow-up period by 21 years and includes a greater number of incident breast cancer cases. The listed life events occurring in the five years prior to assessment are qualified as “stressful” according to the Holmes and Rahe scale ^24^. The increase in statistical power, attributed to the longer follow-up time and, consequently, the number of breast cancer cases, reasserts previous findings and shows that accumulating multiple stressful life events, regardless of their type, significantly increase the risk of breast cancer incidence in the long-term. Leveraging twin pair analyses, we established that the heightened risk of breast cancer is attributable to factors that are not shared between the twins, i.e., the increased risk in breast cancer incidence following stressful life events is independent of genetic background. Prior research has indicated that associations of stressful life events with breast cancer risk are not consistently observed., One explanation for these discrepancies may lie in study design differences, such as variation in the follow-up duration, methods of stress assessment, and inclusion of potential confounding variables. Many prior studies relied on retrospective self-reported stress measures, which are prone to recall bias. Our study mitigates this limitation by using a prospective cohort design with stressful life events data collected decades before cancer diagnosis, reducing the likelihood of recall bias and reverse causation.

Genetic and epigenetic analyses in our study provide valuable insight into the potential biological mechanisms linking stress to breast cancer risk. The PRS analysis indicates that the observed association between stress and breast cancer is unlikely to be due to shared genetic predisposition, as we found no significant association between breast cancer PRS and stressful life event scores. While prior studies have explored PRS in breast cancer risk prediction (Mars et al., 2020) ^4^, few have examined whether genetic susceptibility modifies the impact of psychological stress. In our cohort, the association between breast cancer PRS and incident breast cancer was weak (HR = 1.01–1.23), notably smaller than that reported by Mars et al. (2020) ^4^, which were comparable to other European populations ^25^. The discrepancy may be attributable to differences between the FinnGen cohort used by Mars et al. (2020) ^4^ and our twin cohort, as well as the longer follow-up period in our study, during which environmental risk factors may have exerted a stronger influence, potentially masking genetic contributions. Additionally, while twin studies estimate that approximately 31% of breast cancer risk is attributable to genetic factors ^2^, the PRS model used in this study explains less than 1% of total breast cancer risk ^4^, highlighting inherent limitations in current polygenic models. Although life events are external occurrences, individual responses to stressful life events are influenced by biological and potentially genetic factors ^26–28^. The variability in how individuals perceive, and report stressful experiences may introduce additional complexity in assessing their role in breast cancer risk. However, our findings suggest that genetic predisposition does not confound the stress–breast cancer association, reinforcing the role of environmental exposures in shaping disease risk.

In contrast to the PRS findings, our DNA methylation analysis suggests that stress-related epigenetic modifications may serve as a biological pathway linking stressful life events to breast cancer. We identified 42 CpG sites belonging to 31 genes where DNA methylation changes were associated with both stressful life events and increased breast cancer risk. Importantly, these DNA methylation patterns have been linked to environmental breast cancer risk in our previous study ^20^. To ensure that these associations are not merely a consequence of an underlying undiagnosed cancer, we replicated our findings in a separate dataset of healthy twin pairs, where similar effects of stressful life events on DNA methylation were observed. This suggests that the observed DNA methylation changes are likely independent of a subsequent breast cancer diagnosis.

Prior research supports this connection, as stress exposure has been shown to modify DNA methylation patterns ^9–11,29,30^. Additionally, stress can disrupt endocrine hormone balances ^31,32^, which in turn are known to influence DNA methylation ^33^. This suggests that stressful life events might leave epigenetic marks on DNA either directly or through endocrine-mediated mechanisms, potentially altering the function of breast cancer susceptibility genes and influencing pathways that contribute to tumorigenesis. Notably, these DNA methylation changes were observed, on average, 16 years after stressful life event exposure but up to 11 years before breast cancer diagnosis, reinforcing their potential role in cancer initiation rather than being a consequence of the disease itself.

However, future research is needed to establish a causal relationship between DNA methylation and breast cancer risk, as well as to investigate whether similar epigenetic mechanisms contribute to other cancers with overlapping risk factors or shared driver genes. Functional studies exploring how these DNA methylation changes impact relevant biological pathways, along with analyses of breast tissue-specific DNA methylation, could provide further insight into the mechanisms underlying stress-related cancer susceptibility.

A noteworthy feature of our dataset is that individuals were born across different decades, often prior to other studies referenced in the literature ^16^. While no clear trend in genetic risk across these cohorts was visible when stratifying the data based on decade of birth, we did observe a higher HR for stressful life events in individuals born before 1950. The observed difference in the risk ratio associated with stressful life events may be reflective of the turbulent times in Finnish history, such as the 1930s depression, World War II and the immediate post-war period. Only from the 1950s did economic and social development happen within a long period of peace. Another notable finding is that of oral contraceptive use, whereby women born prior to 1950 had 28% increase in risk for breast cancer as compared to virtually no effect among those born after 1950. This may be due to the changing doses of oral contraceptives, as pills transitioned from high-dose oestrogen formulations (50-100 mcg) in the 1960s to lower doses (10-20mcg).

The epidemiology of breast cancer in Finland is evolving. The cumulative hazard for those born after 1950 in the current cohort was shown to be significantly higher than those born earlier (Supplemental Figure 1), reflecting the general trend of increasing breast cancer incidence in the Finnish population (Finnish Cancer Registry Incidence Figure from www.cancer.fi). However, birth cohort analyses in the entire Finnish population based on the NORDCAN database suggest that the age-specific breast cancer incidence of women born from 1950 onwards is no longer increasing (Supplemental Figure 2, NORDCAN visualisation from https://nordcan.iarc.fr/). The same change can be seen for Sweden as well in the NORDCAN data. Our stratified analysis delineates the importance of timeframe when considering risk associations with breast cancer. There is potentially a paradigm shift in the epidemiology of breast cancer indexed by a major change in incidence for women born in the post-WWII decade in at least two Nordic countries. Our suggestive findings for stressful life events point to greater resilience among women born in the 1950s. These intriguing but preliminary findings warrant further investigation to consolidate the reasons underlying these changes in the epidemiology of breast cancer.

Our study has some limitations. First, while we controlled for multiple confounders, unmeasured psychosocial and behavioural factors may still play a role. Second, DNA methylation data were derived from blood samples, which may not fully reflect epigenetic changes in breast tissue. Future studies should explore tissue-specific DNA methylation patterns to strengthen causal inference. Third, our results are confined to women of European origin, specifically of Finnish descent, highlighting the need for studies in more diverse populations before broader generalizations can be made. Fourth, our study did not include data on high-impact breast cancer variants, such as those located in *BRCA1* and *BRCA2*, which limits a comprehensive examination of their potential influence on breast cancer risk. These variants interact with PRS in a non-additive manner, as demonstrated by Mars et al. (2020) ^4^, and given the number of breast cancer cases in our cohort, any analyses of high-impact genes would be severely constrained. Lastly, one of the challenges we encountered was the limited statistical power to conduct mediation analysis, particularly in evaluating the direct role of DNA methylation as a mediator between stress and breast cancer incidence. Although we observed stress-related DNA methylation changes, our sample size hindered our ability to robustly test whether these changes are truly mediating the observed relationship. In this context, the findings should be interpreted with caution, and further studies with larger sample sizes would be essential to confirm whether DNA methylation truly mediates the stress-breast cancer link. Future work could also benefit from more detailed examination of the functional consequences of these epigenetic changes, which may help clarify the mechanisms at play.

## CONCLUSION

This investigation contributes to our understanding of the delicate aetiology of breast cancer and further solidifies a concerning trend: stressful life events appear to be a major long-acting risk factor for breast cancer. Cumulative stressful life events emerged as a potent risk factor, with a hazard ratio exceeding that of other measured factors across generations, especially for women born prior to 1950. While genetics has an effect, it is likely weak, as our analysis suggests that the majority of breast cancer incidence is likely influenced by non-genetic factors. Additionally, epigenetic changes linked to breast cancer risk were associated with reported experience of stressful life events. Remarkably, these changes persisted on average at least 16 years after the stressful life events and, on average, 11 years before diagnosis. Our findings highlight the critical need to understand the role of stress and stressful life event in breast cancer aetiology.

## DECLARATIONS

### Author Contributions

The study was initiated and designed by Jaakko Kaprio, Elissar Azzi, Hannes Bode and Miina Ollikainen. Data acquisition and preparation were performed by Jaakko Kaprio, Miina Ollikainen, Hannes Bode, Teemu Palviainen and Mikaela Hukkanen. Analysis of the data was conducted by Jaakko Kaprio, Hannes Bode, and Elissar Azzi. The first draft of the manuscript was written by Elissar Azzi and Hannes Bode, and all authors commented on previous versions of the manuscript. All authors read and approved the final manuscript.

### Ethics approval

This study has been approved by the appropriate national research ethics committees, with the most recent approval granted by the Hospital District of Helsinki and Uusimaa ethics board in 2018 (#1799/2017). Permission for linkage to the Finnish Cancer Registry was provided by the Finnish Social and Health Data Permit Authority Findata (THL/6353/14.06.00/2023).

### Consent to participate

Blood samples for DNA analyses were collected from each participant after they had signed a written informed consent.

## Data Availability

The genetic and epigenetic data and associated phenotypes utilized in the current study are stored in the Biobank of the Finnish Institute for Health and Welfare, Helsinki, Finland. The data is publicly available for use by qualified researchers through a standardized application procedure. https://thl.fi/en/web/thl-biobank/forresearchers. The epidemiological analysis data are available through the Institute for Molecular Medicine Finland (FIMM) Data Access Committee (DAC) for authorized researchers who have IRB/ethics approval and an institutionally approved study plan. To ensure the protection of privacy and compliance with national data protection legislation, a data use/transfer agreement is needed, the content and specific clauses of which will depend on the nature of the requested data. Requests will be addressed in a reasonable time frame and the primary mode of data access is by either personal visit or remote access to a secure server.

