## Supplemental Material for "Long-Term Impact of Stressful Life Events on Breast Cancer Risk: A 36-Year Genetically Informed Prospective Study in the Finnish Twin Cohort"

Stressful life events, breast cancer, cohort study, twins, polygenic risk score, methylation

**ORCID:**
Elissar Azzi: 0009-0004-1091-7346
Hannes Frederik Bode: 0000-0003-3232-6518
Teemu Palviainen: 0000-0002-7847-8384
Mikaela Hukkanen: 0000-0002-8574-8274
Miina Ollikainen: 0000-0003-3661-7400
Jaakko Kaprio: [0000-0002-3716-2455](https://orcid.org/0000-0002-3716-2455)

Supplemental Material
This document contains all supplementary tables and figures, as well as in-depth description of analyses mentioned in the manuscript

**Epidemiological Analyses**

**Final sample**

Altogether 563 women (4.3%) did not reply to any items of the life event scale, while 10 820 completed it with at most two missing items (11.2% one item missing and 3.3% with two missing items). After excluding those with prevalent breast cancer or residence outside Finland, 10478 participants remained with valid life event information. Those respondents without valid life event information did not differ in their risk of breast cancer from those included in the analysis (age-adjusted HR=0.83, 95%CI 0.67-1.02).

There was a substantial fraction of missing values for reproductive class (n=459, 4.6%), oral contraceptive use (n=1053, 10%) and social class (n=581, 5.9%), so an additional class of missingness was created for these three categorical variables. Missingness in other covariates was small and the final analysis sample size was 10342, with 719 incident breast cancers from1982 to 2018.

**Supplemental Table 1**
Baseline characteristics reported in 1981 follow-up survey among 10342 women, of whom 719 had incident of breast cancer in the Finnish Twin Cohort, 1982–2018.*

| Risk Factor | Characteristic | Control | Breast cancer cases |
| --- | --- | --- | --- |
| Mean age at response to 1981 survey |  | 39.1 (13.0) | 38.1 (11.0) |
| Mean sum of life events |  | 3.6 (2.5) | 3.9 (2.5) |
| Leisure time physical activity | Lowest | 1580 (16.4%) | 95 (13.2%) |
|  | Low | 1984 (20.6%) | 161 (22.4%) |
|  | Intermediate | 2260 (23.5%) | 199 (27.7%) |
|  | High | 2117 (22.0%) | 157 (21.8%) |
|  | Highest | 1682 (17.5%) | 107 (14.9%) |
| Zygosity | MZ | 2976 (30.7%) | 221 (30.7%) |
|  | DZ | 5902 (61.3%) | 459 (62.9%) |
|  | XZ | 745 (7.7%) | 46 (6.4%) |
| Marital status | Single | 2031 (21.1%) | 162 (22.5%) |
|  | Married | 5963 (62.0%) | 443 (61.6%) |
|  | Remarried | 117 (1.2%) | 10 (1.4%) |
|  | Cohabiting | 531 (5.5%) | 38 (5.3%) |
|  | Divorced | 510 (5.3%) | 39 (5.4%) |
|  | Widowed | 471 (4.9%) | 27 (3.8%) |
| Social class | Upper white-collar | 420 (4.4%) | 39 (5.4%) |
|  | Lower white-collar | 2982 (31.0%) | 261 (36.3%) |
|  | Skilled workers | 2758 (28.7%) | 187 (26.0%) |
|  | Unskilled workers | 1066 (11.1%) | 70 (9.7%) |
|  | Farmers | 689 (7.2%) | 41 (5.7%) |
|  | Others | 1159 (12.0%) | 89 (12.4%) |
|  | Missing data | 549 (5.7%) | 32 (4.5%) |
| Reproductive class  (LCA) | Childless | 3693 (38.4%) | 271 (37.7%) |
|  | 1 | 1082 (11.2%) | 78 (10.8%) |
|  | 2 | 1501 (15.6%) | 108 (15.0%) |
|  | 3 | 1175 (12.2%) | 97 (13.5%) |
|  | 4 | 941 (9.8%) | 80 (11.1%) |
|  | 5 | 647 (6.7%) | 49 (6.8%) |
|  | 6 | 153 (1.6%) | 8 (1.1%) |
|  | Missing data | 431 (4.5%) | 28 (3.9%) |
| Oral contraceptives use | Never user | 3927 (40.8%) | 272 (37.8%) |
|  | Ever user | 4706 (48.9%) | 384 (53.4%) |
|  | Missing data | 990 (10.3%) | 63 (8.8%) |
| Alcohol consumption (grams ethanol per day) | 0 | 3228 (33.5%) | 217 (30.2%) |
|  | 1-10 | 5728 (59.5%) | 447 (62.2%) |
|  | 11-20 | 396 (4.1%) | 32 (4.5%) |
|  | 21 or more | 271 (2.8%) | 23 (3.2%) |
| Smoking status | Never | 5761 (59.9%) | 432 (60.1%) |
|  | Occasional | 220 (2.3%) | 11 (1.5%) |
|  | Former | 1512 (15.7%) | 113 (15.7%) |
|  | Current | 2130 (22.1%) | 163 (22.7%) |
| Mean BMI category | Underweight | 564 (5.9%) | 39 (5.4%) |
|  | Normal | 7142 (74.2%) | 556 (77.3%) |
|  | Overweight | 1580 (16.4%) | 99 (13.8%) |
|  | Obese | 337 (3.5%) | 25 (3.5%) |
| Sum of life events | 0-1 | 2107 (21.9%) | 127 (17.7%) |
|  | 2-3 | 2980 (31.0%) | 231 (32.1%) |
|  | 4-5 | 2391 (24.8%) | 198 (27.5%) |
|  | 6-7 | 1416 (14.7%) | 105 (14.6%) |
|  | 8-9 | 530 (5.5%) | 39 (5.4%) |
|  | 10 or more | 199 (2.1%) | 19 (2.6%) |

*Data is presented as means and standard deviations, or percentages.
 ¶ Sum of life events between 11-17 are collapsed into 10.

XZ, Unknown Zygosity

**Supplemental Table 2**
Hazard ratios derived from univariable and multivariable survival models of known or suspected individual risk factors for breast cancer among 10, 342 women, of whom 719 had incident of breast cancer during 1982–2018.

| Risk Factor | Characteristic | Multivariate HR | 95%CI | Univariate HR | 95%CI |
| --- | --- | --- | --- | --- | --- |
| Zygosity | MZ | 1 |  | 1 |  |
|  | DZ | 1.04 | 0.88-1.24 | 1.03 | 0.87-1.22 |
|  | XZ | 0.95 | 0.67-1.35 | 0.9 | 0.65-1.24 |
| Marital status | Single | 1.33 | 1.09-1.62 | 1.27 | 1.06-1.52 |
|  | Married | 1 |  | 1 |  |
|  | Remarried | 1.11 | 0.60-2.07 | 1.2 | 0.64-2.22 |
|  | Cohabiting | 1.14 | 0.80-1.61 | 1.23 | 0.88-1.73 |
|  | Divorced | 0.98 | 0.70-1.37 | 1.04 | 0.75-1.45 |
|  | Widowed | 1 | 0.67-1.49 | 0.92 | 0.62-1.37 |
| Oral contraceptive use | Never user | 1 |  | 1 |  |
|  | Ever user | 1.19 | 1.00-1.41 | 1.27 | 1.07-1.49 |
|  | Missing data* | 1 | 0.76-1.32 | 0.95 | 0.72-1.25 |
| Social class | Upper white-collar | 1 |  | 1 |  |
|  | Lower white-collar | 1.05 | 0.74-1.47 | 1.05 | 0.75-1.47 |
|  | Skilled workers | 0.89 | 0.62-1.26 | 0.84 | 0.60-1.19 |
|  | Unskilled workers | 0.87 | 0.59-1.29 | 0.81 | 0.55-1.20 |
|  | Farmers | 0.82 | 0.52-1.30 | 0.68 | 0.44-1.06 |
|  | Others | 1.17 | 0.79-1.73 | 1.23 | 0.84-1.79 |
|  | Missing data* | 0.81 | 0.43-1.52 | 0.81 | 0.51-1.31 |
| Reproductive class | Childless | 1 |  | 1 |  |
|  | 1 | 1.15 | 0.89-1.49 | 1.05 | 0.82-1.35 |
|  | 2 | 1.1 | 0.87-1.40 | 1 | 0.80-1.25 |
|  | 3 | 1.24 | 0.97-1.59 | 1.17 | 0.92-1.49 |
|  | 4 | 1.3 | 1.00-1.69 | 1.27 | 0.98-1.63 |
|  | 5 | 1.2 | 0.88-1.64 | 1.21 | 0.89-1.65 |
|  | 6 | 0.95 | 0.46-1.96 | 0.74 | 0.37-1.50 |
|  | Missing data‡ | 1.26 | 0.71-2.25 | 0.98 | 0.65-1.46 |
| BMI class | Underweight | 0.97 | 0.70-1.35 | 1.07 | 0.77-1.48 |
|  | Normal | 1 |  | 1 |  |
|  | Overweight | 0.8 | 0.64-1.00 | 0.72 | 0.58-0.90 |
|  | Obese | 1.06 | 0.70-1.60 | 0.93 | 0.62-1.39 |
| Alcohol consumption  (ethanol gram per day) | 0 | 1 |  | 1 |  |
|  | 1-10 | 1.07 | 0.89-1.29 | 1.21 | 1.02-1.44 |
|  | 11-20 | 1.10 | 0.73-1.67 | 1.32 | 0.89-1.96 |
|  | 21 or more | 1.24 | 0.79-1.94 | 1.48 | 0.96-2.27 |
| Smoking | Never | 1 |  | 1 |  |
|  | Occasional | 0.65 | 0.35-1.18 | 0.73 | 0.40-1.32 |
|  | Former | 1 | 0.80-1.26 | 1.11 | 0.89-1.38 |
|  | Current | 1.08 | 0.88-1.33 | 1.21 | 1.00-1.46 |
| Leisure time physical activity quintiles | Lowest | 1 |  | 1 |  |
|  | Low | 1.22 | 0.94-1.58 | 1.3 | 1.01-1.68 |
|  | Intermediate | 1.27 | 0.99-1.63 | 1.4 | 1.10-1.79 |
|  | High | 1.06 | 0.81-1.38 | 1.18 | 0.91-1.52 |
|  | Highest | 0.92 | 0.69-1.21 | 1.02 | 0.78-1.35 |

Same number of cases and person-time in all analyses
Multivariate HRs are adjusted for the influence of other covariates included in the model: zygosity, marital status, oral contraceptive use, social class, reproductive class, BMI, alcohol consumption, smoking, leisure time physical activity.
HR, Hazard ratio; CI, Confidence interval, XZ, Unknown Zygosity

**Multivariable-adjusted survival models of individual risk factors for breast cancer stratified by birth cohort**

The ever (current or former) use of oral contraceptives was shown to increase HR to 1.27 (95%CI 1.07-1.49) as compared to never-users. After adjustment for other potentially confounding factors, the HR decreased slightly to 1.19 (95%CI 1.00-1.41). Marital status, specifically being single, was shown to be significantly associated with a heightened breast cancer risk, with an HR between 1.27 and 1.33 (univariable and multivariable analysis, 95%CI 1.06-1.52 and 1.09-1.62), while other marital status categories were not associated. Compared to never smokers, current smokers had an HR of 1.21 (95%CI 1.00-1.46). This association was attenuated in the multivariable analysis with an HR of 1.08 (95%CI 0.88-1.33). The hazard ratios associated with alcohol consumption increased with the amount of ethanol ingested per day, whereby 1-10 grams of ethanol per day augments HR to 1.21 (95%CI 1.02-1.44), and a positive trend of higher point estimate of risk with higher daily consumption of ethanol: 1. 32 (95%CI 0.89-1.96) for 11-20 g/day and 1.48 (95%CI 0.96-2.27) for 21 or more grams of ethanol per day. The risk estimates for alcohol consumption attenuated when controlling for confounding factors, but the trend remains consistent. Interestingly, low and intermediate leisure time (2^nd^ and 3^rd^ quintiles) physical activity levels are associated with an increased estimate, 1.3 (95%CI 1.01-1.68) and 1.4 (95%CI 1.10-1.97) respectively, as compared to the lowest quintile of activity level. Multivariable adjustment slightly decreases HR estimates. Analysis of BMI indicated that overweight showed a decreased HR of 0.72 (95%CI 0.58-0.90) as compared to normal BMI. Zygosity, social class and reproductive class present no significant associations.

**Supplemental Table 3**
Multivariable-adjusted HR of known or suspected individual risk factors for breast cancer stratified by birth cohort among 10, 342 women, of whom 719 had incident of breast cancer during 1982–2018.

| Risk factor | Characteristic | All | 95%CI | Before 1950 | 95%CI | After 1950 | 95%CI |
| --- | --- | --- | --- | --- | --- | --- | --- |
| Stressful life events |  | 1.04 | 1.00-1.07 | 1.05 | 1.01-1.10 | 1 | 0.95-1.05 |
| Marital status | Single | 1.33 | 1.09-1.62 | 1.18 | 0.89-1.58 | 1.31 | 0.93-1.83 |
|  | Married | 1 |  | 1 |  | 1 |  |
|  | Remarried | 1.12 | 0.60-2.08 | 1.36 | 0.71-2.62 | 0.47 | 0.06-3.39 |
|  | Cohabiting | 1.14 | 0.80-1.61 | 1.29 | 0.76-2.20 | 0.97 | 0.60-1.58 |
|  | Divorced | 0.98 | 0.70-1.37 | 0.97 | 0.66-1.43 | 1.22 | 0.61-2.45 |
|  | Widowed | 1 | 0.67-1.50 | 0.99 | 0.65-1.51 | 4.33 | 1.08-17.41 |
| Oral contraceptive use | Never user | 1 |  | 1 |  | 1 |  |
|  | Ever user | 1.19 | 1.00-1.41 | 1.28 | 1.03-1.59 | 1.01 | 0.75-1.35 |
|  | Missing data | 1 | 0.76-1.32 | 1.02 | 0.74-1.41 | 1.09 | 0.60-1.99 |
| BMI class | Underweight | 0.97 | 0.70-1.35 | 0.97 | 0.53-1.77 | 0.91 | 0.61-1.36 |
|  | Normal | 1 |  | 1 |  | 1 |  |
|  | Overweight | 0.8 | 0.64-1.00 | 0.82 | 0.64-1.05 | 0.9 | 0.50-1.61 |
|  | Obese | 1.05 | 0.70-1.60 | 1.2 | 0.79-1.85 | 0 | 0.00-0.00 |
| Alcohol consumption (ethanol grams per day) | 0 | 1 |  | 1 |  | 1 |  |
|  | 1-10 | 1.07 | 0.89-1.29 | 1.14 | 0.91-1.43 | 0.89 | 0.64-1.25 |
|  | 11-20 | 1.1 | 0.73-1.67 | 1.1 | 0.65-1.89 | 1.05 | 0.54-2.05 |
|  | 21-30 | 1.24 | 0.79-1.94 | 1.07 | 0.57-1.99 | 1.37 | 0.71-2.62 |
| Smoking | Never | 1 |  | 1 |  | 1 |  |
|  | Occasional | 0.65 | 0.35-1.18 | 0.68 | 0.32-1.44 | 0.62 | 0.22-1.71 |
|  | Former | 1 | 0.80-1.26 | 1.08 | 0.81-1.44 | 0.89 | 0.62-1.29 |
|  | Current | 1.08 | 0.88-1.33 | 0.91 | 0.68-1.21 | 1.25 | 0.91-1.71 |
| Leisure time physical activity | Lowest | 1 |  | 1 |  | 1 |  |
|  | Low | 1.22 | 0.94-1.58 | 1.2 | 0.87-1.66 | 1.22 | 0.79-1.90 |
|  | Intermediate | 1.27 | 0.99-1.63 | 1.35 | 0.99-1.84 | 1.14 | 0.75-1.74 |
|  | High | 1.06 | 0.81-1.38 | 1.15 | 0.83-1.60 | 0.93 | 0.59-1.47 |
|  | Highest | 0.92 | 0.69-1.21 | 0.82 | 0.57-1.18 | 1.07 | 0.69-1.67 |
| Time at risk (person-years) |  | 332906 |  | 194876 |  | 138031 |  |
| Number of breast cancer cases |  | 719 |  | 452 |  | 267 |  |
| Number of persons |  | 10342 |  | 6398 |  | 3944 |  |

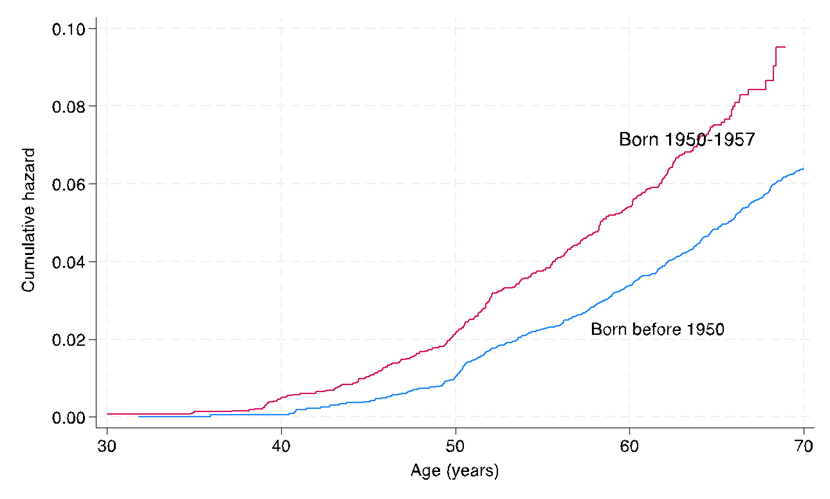

**Supplemental Figure 1: Breast cancer incidence by birth cohort among 10342 women, of whom 719 had incident of breast cancer in the Finnish Twin Cohort during 1982–2018**

This figure displays the age-dependent breast cancer incidence rates among 10342 women enrolled in the Finnish Twin Cohort, categorized by birth cohorts born before 1950 and from 1950 to 1957

- This Figure was created with Stata version 18.

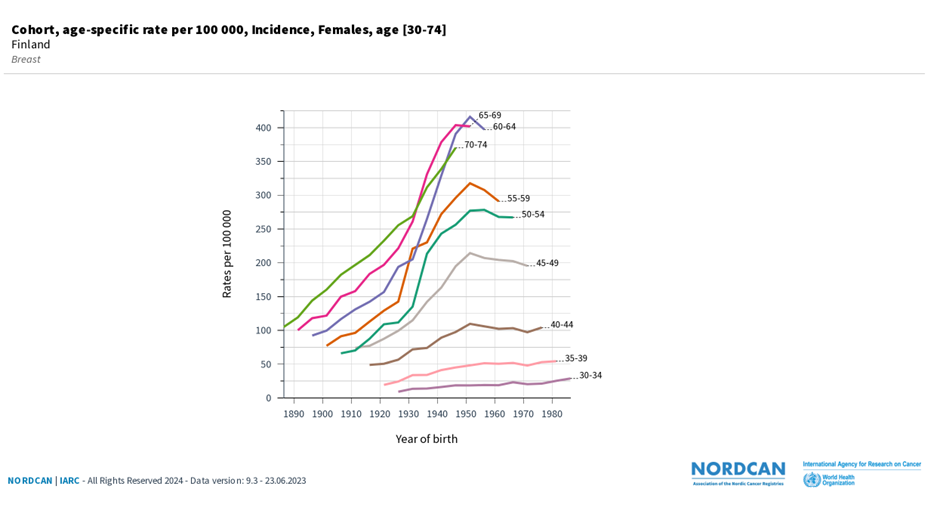

**Supplemental Figure 2: Age-specific breast cancer incidence rates per 100000 females in Finland**

This figure displays the age-specific breast cancer incidence rates per 100000 females in Finland born across different decades as obtained from the NORDCAN visualization platform. Source: Association of the Nordic Cancer Registries (ANCR) & International Agency for Research on Cancer (IARC). Available from<https://nordcan.iarc.fr/> (Accessed: 20/04/2024)

- This Figure was created with the Nordcan visualization platform.

**Genetic Analyses**

**Polygenic Risk Score for Breast Cancer**

**Cohort description for polygenic risk score analysis**
Genetic material for evaluating genetic breast cancer risk through PRS was accessible for 4601 individuals (4237 controls and 364 cases, disease cumulative incidence 7.9%). Within this set of women, we identified DZ sister pairs discordant for breast cancer (176 cases with breast cancer diagnosis after the 1981 questionnaire and their 176 sisters). Given the same genomic sequence in MZ pairs, PRS analyses in such pairs was not undertaken. In addition, there were 17 DZ twin pairs and 15 MZ twin pairs concordant for breast cancer during the follow-up period.
To explore the impact of breast cancer risk factors on the relationship between risk of breast cancer and breast cancer PRS, the dataset was narrowed down to individuals with complete data on these factors including exposure to stressful life events, resulting in 3906 women (3585 controls and 321 cases, prevalence 8.2%); among them 141 DZ twin pairs discordant for breast cancer.

**Genetic data and polygenic risk score generation**
A breast cancer PRS was derived from GWAS summary statistics of breast cancer by Michailidou et al. (2017) ^19^. The total number of single nucleotide polymorphisms used for the PRS calculation were 1069529. The number of women in the original GWAS study was 228,951. The calculation of the PRS involved the utilisation of HapMap3 SNPs with a European Minor Allele Frequency (MAF) exceeding 5%. To improve accuracy, a cohort comprising 27,284 individuals from the FINRISK dataset was incorporated as a Linkage Disequilibrium (LD) reference panel. The technical details of genotyping, imputation and PRS calculation have been described elsewhere ^34^. The 1069529 SNPs explain about 0.82% variability in breast cancer risk. Subsequently, samples that did not pass the quality control and those that originated from pairs that showed deviations in their expected genetic share (between 1.00 and 0.95 for MZ twin pairs and between 0.65 and 0.35 for DZ twin pairs) were excluded from the analysis.

Population genetic variation was corrected by linearly regressing the breast cancer PRS over the first 10 Principal Components (PCs) of the genotype data. Subsequently the residuals were used for the analysis. Following this correction, the PRS data for the 4601 women was scaled on the standard deviation and mean-centred.

**Survival analysis for genetic breast cancer risk**

A survival analysis was performed to investigate the association between the breast cancer PRS and breast cancer incidence. This analysis incorporated the twin pair relationship to account for shared familial factors. The Cox Proportional Hazards model within the R package *survival* ^35^ was used. The proportional hazards assumption was assessed using the cox.zph() function and visual inspection of Schoenfeld residuals. This analysis mirrored the above performed overall phenotype analysis.

Two models were fitted to explore the association between breast cancer PRS and breast cancer incidence. The first model had the largest sample size as it included all available individuals. The second model, a co-twin control model, focused only on discordant DZ twin pairs, which controls for shared familial effects. Within the co-twin control model, a sub-model was fitted to further explore the separate contributions of the pairwise mean PRS and the within-pair PRS difference. This allows for a more nuanced understanding of how PRS influences breast cancer risk within families.

To assess the robustness of the findings, sensitivity analyses were conducted where several additional survival models were fitted. First, individual and discordant pair models were fitted for individuals with complete phenotypic data. These models were run both unadjusted and adjusted for the available breast cancer risk factors. This allows for comparison of the PRS effect with and without the influence of other risk factors and can determine whether the PRS is independent of such. Finally, individual models were stratified by the above defined birth cohort to assess potential variations in the PRS effect across different generations.

**Results based on Breast Cancer PRS and cancer incidence**

To investigate the association between the breast cancer PRS and breast cancer incidence, multiple survival analyses were performed (Supplemental Table 4). The initial analysis included 4237 female controls and 364 breast cancer cases. This analysis adjusted for underlying family structure and found a 3% increase in breast cancer risk per SD increase in breast cancer PRS, with a HR of 1.03 (95%CI 0.92-1.15). To further account for familial effects and potential unknown confounders, a subsequent analysis focused specifically on 176 DZ twin sister pairs discordant for breast cancer. This model yielded a similar HR of 1.03 (95%CI 0.89-1.20). However, a more nuanced analysis was subsequently conducted for the discordant DZ twin pairs, incorporating both the within-pair mean breast cancer PRS and the within-pair differences in breast cancer PRS separately. Here, the within-pair mean PRS was not associated with breast cancer risk. However, the within-pair difference in breast cancer PRS showed a greater point estimate of risk, with an HR of 1.20 (95%CI 0.88-1.62). This suggests that the relative difference in PRS between twins within a pair may be a more informative indicator of risk compared to the absolute PRS value. This finding supports the notion that individuals with higher breast cancer PRS scores have an increased risk of breast cancer, even if the increase is modest in the present data set.

To assess data robustness, we conducted additional analyses. At first, we compared observed within-pair breast cancer PRS with expected values (0.5 for DZ twins, 1.0 for MZ twins). DZ twin pairs discordant for breast cancer did not deviate from their expected correlation (Supplementary Figure 3). Only concordant DZ twin pairs deviated from the expected value of 0.5. DZ pairs concordant for breast cancer showed a within pair correlation of 0.63 (95%CI 0.22-0.85, n=17 pairs) for breast cancer PRS, while healthy concordant DZ pairs showed a within pair correlation of 0.53 (95%CI 0.49-0.57, n=1035 pairs), however these observations were not statistically significant (Supplementary Figure 3). We explored potential variations in genetic breast cancer risk across birth cohorts. Analyses revealed a non-significant trend towards higher risk for women born after 1940 compared to those born before (Supplemental Table 4). However, confidence intervals overlapped across birth cohorts, suggesting inconclusive results. Further analyses showed that the collected phenotype data had no significant impact on genetic risk estimates (Supplemental Table 4). Only, participation in the genetic study part was linked to a slightly higher breast cancer risk (OR 1.23, 95%CI 1.07-1.41).

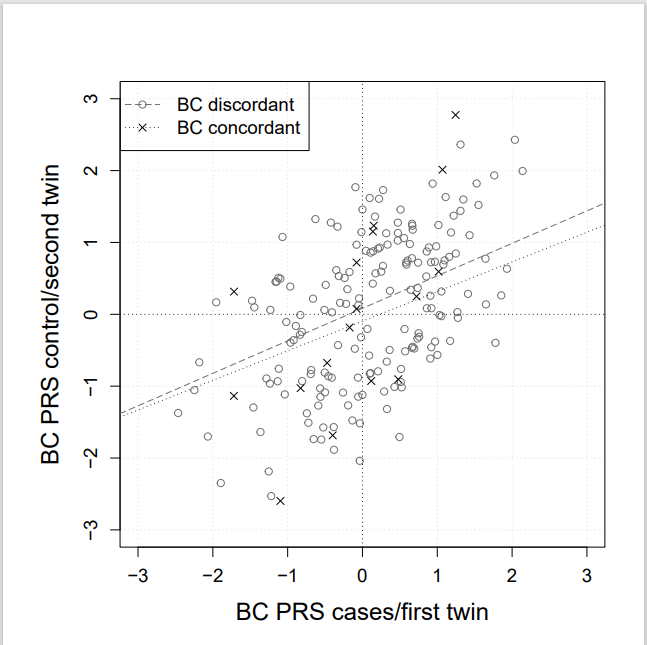

**Supplemental Figure 3: Pairwise comparison of breast cancer PRS values between 176 breast cancer discordant and 17 concordant dizygotic twin pairs**

The x-axis represents the breast cancer (BC) PRS for the cases or twin diagnosed with breast cancer first during the 36 years of follow-up, while the y-axis represents the breast cancer (BC) PRS for the controls or twin diagnosed second. The figure contains trend lines for both discordant and concordant twin pairs.

- This Figure was created with R version 4.3.2.

**Supplemental Table 4: Survival analysis of breast cancer PRS: Individual and breast cancer discordant twin pair models, adjusted and unadjusted for epidemiological risk factors, stratified by birth cohort**

| **Model** | **Cov. adj.** | **Controls** | **Cases** | **HR (95%CI)**  **breast cancer PRS** | **p** |
| --- | --- | --- | --- | --- | --- |
| Individual model | NO | 4237 | 364 | 1.03 (0.92-1.15) | 0.65 |
| Paired model* | NO | 176 | 176 | 1.03 (0.89-1.2) | 0.66 |
| Paired model*  (Pairwise mean and difference in breast cancer PRS separately) | NO | 176 | 176 | mean: 0.99 (0.83-1.17) | 0.86 |
|  |  |  |  | diff.: 1.2 (0.88-1.62) | 0.25 |
| **Sensitivity analysis** | | | | | |
| Individual model full epi-data | NO | 3585 | 321 | 1.02 (0.90-1.15) | 0.75 |
|  | YES |  |  | 1.01 (0.90-1.15) | 0.82 |
| Paired model full epi-data* | NO | 141 | 141 | 1.05 (0.89-1.24) | 0.55 |
|  | YES |  |  | 1.05 (0.88-1.26) | 0.59 |
| Paired model*  (Pairwise mean and difference in breast cancer PRS separately) full epi-data | NO | 141 | 141 | mean: 1.00 (0.83-1.21) | 0.97 |
|  |  |  |  | diff.: 1.23 (0.87-1.74) | 0.25 |
|  | YES |  |  | mean: 1.00 (0.81-1.23) | 0.99 |
|  |  |  |  | diff.: 1.22 (0.84-1.76) | 0.29 |
| **Per birth decade** | | | | | |
| Before 1930s | NO | 1856 | 158 | 0.90 (0.76-1.08) | 0.26 |
| 1940s | NO | 1379 | 125 | 1.21 (1.00-1.47) | 0.05 |
| 1950s | NO | 1002 | 81 | 1.05 (0.80-1.37) | 0.73 |
| **Before and after 1950** | | | | | |
| Before 1950 | NO | 3235 | 283 | 1.03 (0.90-1.16) | 0.70 |
| After 1950 | NO | 1002 | 81 | 1.05 (0.80-1.37) | 0.73 |

HR, Hazard Ratio; CI, confidence interval; Cov. adj., adjusted for covariates

*, Zygosity not included as only DZ twins

**DNA Methylation Analyses**

**Environmental Breast Cancer Risk related DNA methylation**

To investigate the association between DNAm at the same set of 212 CpG sites previously linked to breast cancer risk in Bode et al. (2024) ^20^ and exposure to stressful life events we performed a set of within-pair analyses.

Here we examined a subsample of 319 twin pairs from the older Finnish Twin Cohort (172 MZ and 147 DZ twin pairs). The average age of the participants at the 1981 questionnaire was 40.20 (SD 9.8) years. The mean age at blood sampling for this cohort was 63.2 (SD 8.3) years. On average twins in a pair had by 3.7 (SD 2.1) stressful life events reported. The MZ twins alone had similar values, with mean age at the 1981 questionnaire of 38.8 (SD 9.8) years, mean age at blood sampling was 64.4 (SD 7.9) years and a mean of 3.9 (SD 2.1) exposures to stressful life events.

For the DNAm data preprocessing we followed a standard protocol described in detail by Bode et al. (2024)^20^. Briefly, this protocol involved quality control checks on raw DNAm data, functional normalisation to remove initial technical variations, beta-mixture quantile normalisation to address specific within-sample probe bias, and scaling of DNAm values for each probe (CpG site) across all samples to eliminate between-sample technical variations. All samples were generated using the Illumina Infinium MethylationEPIC (EPIC) platform (Illumina, San Diego, CA, USA). CpG site annotation was performed using the latest version of the Illumina Infinium Methylation EPIC manifest v1.0 B5 (Illumina, San Diego, CA, USA).

Two similar linear models were employed to analyse the association between within pair difference in DNAm and the within pair difference in stressful life events. The first model included all twin pairs including both MZ and DZ. This approach offered a larger sample size and controlled for familial factors, age, and technical confounders. However, this model does not fully account for germline genetics. Still, due to their high genetic similarity even in DZ twin pairs, most DNAm differences within each pair are likely due to environmental factors influencing breast cancer risk. We regarded nominal significance (p<0.05) and effect direction the same as reported by Bode et al. (2024) ^20^ as meaningful replication.

To strengthen the evidence from the first model, a second model was employed that focused exclusively on the MZ twin pairs with DNAm and SLE data. This approach ensured complete control for germline genetics, but it also resulted in a smaller sample size. Therefore, this second model served to confirm whether the effect sizes observed in the first model remained consistent after accounting for germline genetics.

To compare results by zygosity, we also ran the analysis in DZ pairs alone. This followed the similar approach as in the other analysis. The DZ twins alone had slightly different descriptive values compared to the MZ twins but still in a similar range. The mean age at the 1981 questionnaire was 41.5 (SD 9.6) years, mean age at blood sampling was 61.9 (SD 8.5) years and the mean of exposures to stressful life events was on average 3.4 life events (SD 2.0)

To exclude any confounding by future breast cancer diagnosis independently associating with stressful life events exposure and DNAm signature, we performed a sensitivity analysis using DNAm data from 154 Finnish Twin Cohort female twin pairs (96 MZ and 58 DZ) without any cancer diagnosis until the end of the follow-up in 2018. The average age of the twins at the time of a questionnaire administered in 1981 was 39.2 (SD 9.6) years. The mean age at blood sampling for the DNAm data generation was 65.9 (SD 5.7) years. On average twins in the pairs had exposure to 3.9 (SD 2.0) stressful life events. We first performed the within-pair analyses in the full set of 154 twin pairs, and then restricted the analysis to the 96 MZ twin pairs, as described above.
